# Real-world monitoring of BNT162b2 vaccine-induced SARS-CoV-2 B and T cell immunity in naive healthcare workers: a prospective single center study

**DOI:** 10.1101/2022.01.17.22269081

**Authors:** Bas Calcoen, Kim Callebaut, Aline Vandenbulcke, Nico Callewaert, Xavier Bossuyt, Johan Van Weyenbergh, Piet Maes, Maya Imbrechts, Thomas Vercruysse, Hendrik Jan Thibaut, Dorinja Zapf, Kersten Dieckmann, Karen Vanhoorelbeke, Nick Geukens, Simon De Meyer, Wim Maes

## Abstract

**Background:** Severe acute respiratory syndrome coronavirus 2 (SARS-CoV-2) is the cause of the ongoing COVID-19 pandemic. To prevent the massive COVID-19 burden, several vaccination campaigns were initiated. We performed a single center observational trial to evaluate adaptive immunity in naive healthcare workers upon BNT162b2 vaccination.

**Methods:** Serological analysis was performed through conventional immunoassays. Antibody functionality was analyzed via in vitro neutralization assays. Circulating receptor-binding domain (RBD) specific B cells were assessed via flowcytometry. The induction of SARS-CoV-2 specific T cells was investigated through interferon-γ release assay combined with flowcytometric profiling of activated CD4 and CD8 T cells.

**Results:** Three months after vaccination, all but one of the subjects (N = 31) displayed vaccine-induced neutralizing antibodies. In 10 out of 31 subjects, circulating RBD specific B cells were found of which the rate showed moderate correlation to serological parameters. Specific interferon-γ release was present in all subjects and correlated with the significant upregulation of CD69 on CD4+ and CD8+ T cells and CD40L on CD4+ T cells. Interestingly, no relation was found between B and T cell parameters. In addition, one symptomatic breakthrough infection with the SARS-CoV-2 alpha variant of concern was reported.

**Conclusion:** Three months post vaccination, both humoral and cellular immune responses are detectable in all but one participant. No correlation was found between the magnitude of both B and T cell responses.

## 1 Introduction

Back in December 2019, multiple cases with serious pneumonia of unknown origin (later renamed as coronavirus disease 19 or COVID-19) were described in Wuhan, China (1). Not long afterwards, the causative pathogen was identified via real-time polymerase chain reaction (RT-PCR) as a novel sarbecovirus (sub-genus of the β-coronaviridae) and named severe acute respiratory syndrome coronavirus 2 (SARS-CoV-2) (2,3).

Within a few weeks, SARS-CoV-2 rapidly spread internationally and on 30 January 2020, the world health organization (WHO) declared the outbreak a public health emergency of international concern (4). As a result, many countries introduced strict socio-economic rules (e.g. lockdown, mouth mask) to limit viral spreading and prevent a total collapse of their healthcare system (5).

During the pandemic, multiple strategies including the development of a SARS-CoV-2 vaccine were urgently initiated to lower the massive SARS-CoV-2 related burden. Multiple vaccine development strategies were explored, including nucleic acid-based (e.g. BNT162b2 or Comirnaty^®^ (6,7), mRNA-1732 or Spikevax^®^ (8,9)), adenovector-based (e.g. Vaxzevria^®^ (10), COVID19 vaccine Janssen^®^ (11,12)) and protein-based (Sanofi-GSK^®^ (13)) vaccines of which several products received conditional authorization by either the European Medical Agency (EMA) or Food and Drug Administration (FDA) at the end of 2020. This resulted in the roll-out of international vaccination campaigns at an unprecedented scale and speed.

However, due to the emergence of several SARS-CoV-2 variants of concern (VoC) that already led to new infection waves around the globe (14,15), there is reasonable concern whether the immune response evoked by the currently available vaccines is sufficient to cover these VoC. For example, it is known that several of these SARS-CoV-2 VoC have mutations within their spike (S) protein or receptor-binding domain (RBD) region and therefore could be able to escape functionally active vaccine-induced antibodies that only bind to unique epitopes present on Wuhan strain S or RBD (16). In addition, only a limited number of studies reveal mid-term (i.e. longer than 2 months post mRNA vaccination) sustainability data of vaccine-induced de novo responses to SARS-CoV-2 within healthy subjects (17,18). Finally, most vaccine monitoring studies are often solely based on the evaluation of serological responses (19), although it is known from for example vaccination against smallpox that specific T cells can be detected up to 75 years after vaccination and thus are crucial for long-term immunity (20,21). Indeed, few groups have looked at both vaccine-induced B and T cell immunity, in particular in healthy SARS-CoV-2-naive volunteers (22,23) as most trials focused on either immunocompromised or SARS-CoV-2 convalescent patients. Concerning specific T cell responses upon mRNA-based SARS-CoV-2 vaccination, available data are limited to phase I/II studies (24) of the Comirnaty^®^ vaccine and studies that either assessed short-term T cell response (i.e. up to one month after booster vaccination) (25,26) or focused on T cell responses upon prime or boost vaccination (27,28). Of note, there are several, sometimes contradictory reports concerning both the occurrence rate (29,30) and severity (31,32) of post mRNA vaccination breakthrough infections (BTI).

To address these gaps, we monitored vaccine elicited B and T cell responses in a real-world setting. Hereto, SARS-CoV-2 naive healthcare workers were followed for three months after vaccination with the BNT162b2 SARS-CoV-2 vaccine (Comirnaty^®^) in a prospective single center study.

## 2 Materials and Methods

### 2.1 Study design

#### 2.1.1 Recruitment

During January 2021, professional healthcare workers from the supraregional AZ Groeninge hospital (Kortrijk, Belgium) qualified to receive the BNT162b2 (Comirnaty^®^) vaccine were contacted via the hospital’s newsletter for voluntary enrollment in this study. Healthcare workers were included after signing an informed consent form. Participants were excluded if they met at least one of the following criteria: severe acute infection of any kind, pregnancy, primary immunodeficiency (PID), chronic treatment with immunomodulatory agents (e.g. anti-TNF-α agents, corticosteroids), documented earlier natural infection with SARS-CoV-2 or positive serology (including either anti-S IgG, anti-S IgA or anti-nucleocapsid (N) IgG) found at baseline sampling.

#### 2.1.2 Sampling

A baseline sampling moment prior to vaccination was performed either one day before or on the day of prime vaccination (t_pre_). Comirnaty^®^ administration was performed as indicated by the manufacturer and upon the recommendation by the Belgian Superior Health Council. A second sampling was performed three months after t_pre_ (t_3m_). In case a participant, after receiving two vaccines, presented with COVID-19 symptoms (e.g. dry cough, fever and/or headache) and was positive for SARS-CoV-2 via real-time polymerase chain reaction (RT-PCR), he/she was considered to experience a symptomatic BTI. In this situation, additional sampling was scheduled as soon as possible (t_BTI_). Sampling details are visualized in **Figure 1**.

**Figure 1.**
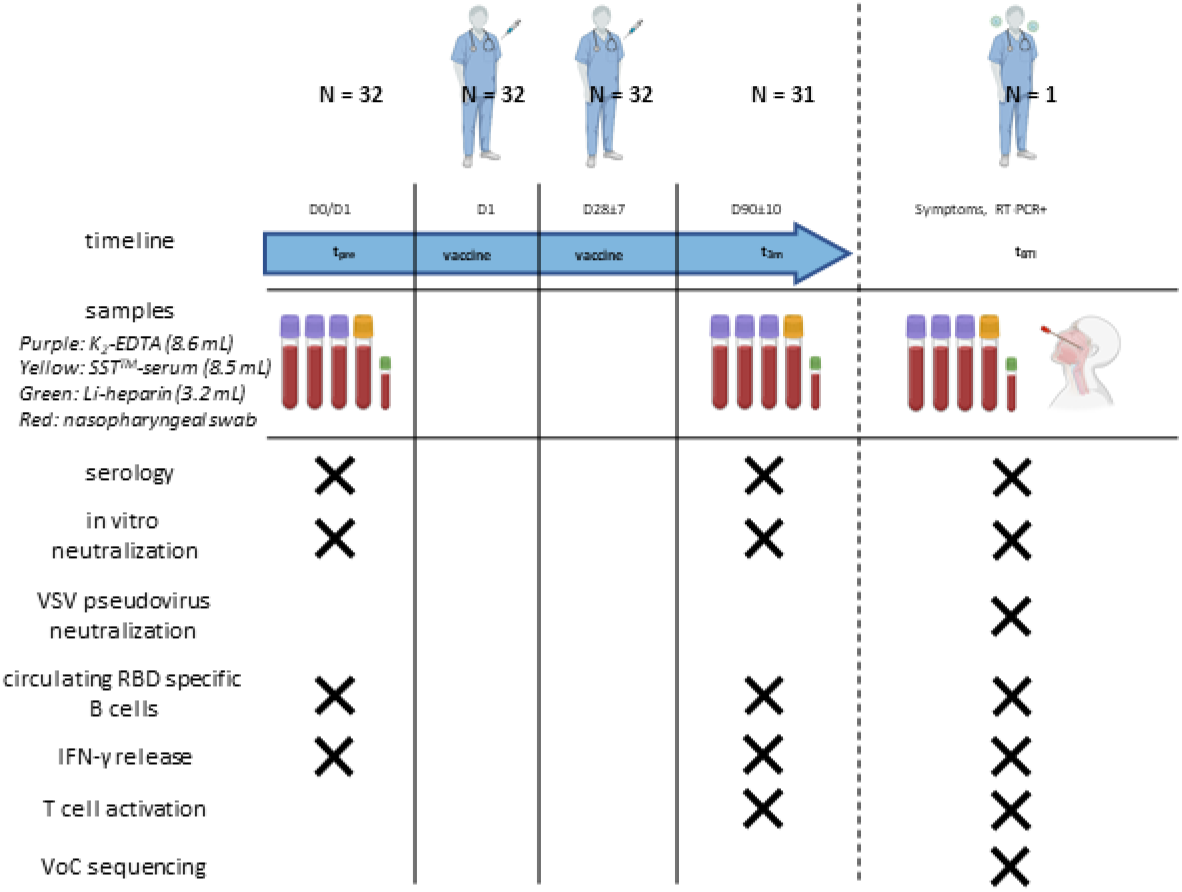
Study design and timeline. A total of 32 healthcare workers were enrolled. One day before or on the day of the first BNT162b2 vaccine, a first baseline sampling moment was scheduled (t_pre_). A second sampling moment (t_3m_) was planned between 80-100 days after baseline.

On each moment, serum was collected to determine SARS-CoV-2 serology and neutralization efficacy whereas blood was used both for isolating PBMCs to detect circulating RBD specific B cells and to assess both SARS-CoV-2 specific T cell activity and IFN-γ release. When a participant developed symptoms and was tested positive via RT-PCR after receiving two vaccines, an additional sampling moment was planned (t_BTI_). At t_BTI_, a nasopharyngeal swab was collected to execute viral whole genome sequencing. Also, a VSV pseudovirus neutralization assay was additionally performed at this timepoint. Abbreviations: PBMC = peripheral blood mononuclear cells, IFN-γ = interferon γ, RT-PCR = real-time polymerase chain reaction, VSV = vesicular stomatitis virus, VoC = variants of concern. This figure was created using Biorender (www.biorender.com).

#### 2.1.3 Serum and peripheral blood mononuclear cell (PBMC) isolation

Venous blood from three K_2_-EDTA tubes was used to isolate PBMCs via density gradient isolation with Ficoll (lymphoprep^™^, STEMCELL technologies, Norway) using SepMate tubes (STEMCELL technologies, Norway). PBMCs were diluted in 90 % fetal calf serum containing 10 % DMSO, divided in aliquots and stored in liquid nitrogen after a controlled cooling procedure. When needed, PBMC aliquots were shortly thawed at 37 °C and washed two times in dPBS (Gibco^™^ Life Sciences, Thermo Fisher Scientific, USA) with a centrifugation step at 300 g for 7 min at 4 °C after each washing. Serum was collected from the SST^™^ II Advance tubes and stored at -20 °C.

#### 2.1.4 Ethical approval

This clinical trial was registered at the EU Clinical Trial Register with ID 2021-001304-15. This trial was performed according to the declaration of Helsinki and was approved by both the local Ethical Committee of the AZ Groeninge hospital (B3962021000022) and the Belgium Federal Agency of Drugs and Health Products (FAGG; protocol no. AZGS2021005).

### 2.2 Serological parameters

#### 2.2.1 Anti-S IgA and IgG assay

Serum anti-S IgA antibodies were measured with the Anti-SARS-CoV-2 IgA enzyme immunoassay from EUROIMMUN (Lübeck, Germany) on an ETI-Max 3000 instrument from DiaSorin (Saluggia, Italy). Following the instructions from the manufacturer, samples with a cut-off index greater or equal to 1.1 were labeled as positive. Serum anti-S IgG titers were measured with the VIDAS SARS-COV-2 IgG (9COG) enzyme immunoassay from Biomérieux (Marcy-l’Etoile, France) on a VIDAS 3 instrument from the same manufacturer. According to the manufacturer’s instructions, samples with a cut-off index greater or equal to 1.0 were considered positive.

#### 2.2.2 Anti-RBD IgG assay

Enzyme-linked immunosorbent assay (ELISA) plates (Corning Costar; cat. Nr. 3590) were coated overnight with His_6_-tagged RBD. Plates were blocked for 2 h at room temperature (RT) using blocking buffer (PBS + 1 % BSA) and washed (wash buffer PBS + 0.002 % Tween 80) six times. Monoclonal anti-RBD calibrator antibody (Sino Biologicals; cat. Nr. 40150-D004) and serum samples (diluted minimum 500-fold) were incubated for 2 h at RT (buffer PBS + 0.1 % BSA + 0.002 % Tween 80). After an additional washing step, goat antihuman (GAH) IgG conjugated with horseradish peroxidase (HRP) was added (1:5000 dilution) and incubated for 1 h at RT. Subsequently, washing was performed and the plate was developed using o-phenylenediamine dihydrochloride (OPD; 0.4 g/L) and H_2_O_2_ (0.003 %) in citrate buffer. After 30 min, the reaction was stopped with H_2_SO_4_ (4 M). The absorbance was measured at 492 nm and the dose-response curve was analyzed by non-linear regression using GraphPad Prism 9.0.0 (Graph Software, San Diego, CA, USA). The assay was validated by measuring assay cut-off values for detection and quantification, accuracy, imprecision and dilutional linearity (33). Sample concentrations were determined using the anti-RBD calibrator antibody and WHO International Standard Serum (NIBSC ref 20-136) (34).

#### 2.2.3 Anti-N IgG assay

The presence of serum anti-N IgG antibodies was determined via the Anti-SARS-CoV-2-NCP (IgG) enzyme immunoassay from EUROIMMUN on an ETI-Max 3000 instrument from DiaSorin. Samples that had a cut-of index greater or equal to 1.1 were considered positive as recommended by the instructions from the manufacturer.

### 2.3 Antibody functionality: neutralization assay

#### 2.3.1 In vitro neutralization assay

Determination of the neutralizing capacity of the vaccine-induced antibodies was generated using the EUROIMMUN SARS-CoV-2 NeutraLISA assay according to the manufacturer’s instructions. Samples were diluted 1:5 in sample buffer. A photometric measurement was made on a wavelength of 450 nm together with a reference wavelength of 620 nm. Semiquantitative results were generated by calculating a ratio of the extinction values of the sample over the mean extinction value of the blank (measured in duplicate) and were presented as percentage inhibition. Percentage inhibition values lower than 20 were defined as negative, between 20 and 35 as borderline and higher or equal to 35 as positive. Lot-specific control concentrates (positive and negative) were used as assay references.

#### 2.3.2 Vesicular stomatitis virus (VSV) pseudovirus neutralization assay

VSV S-pseudotypes were generated as described previously (35). Briefly, HEK-293T cells (SARS-CoV-2) were transfected with the respective S protein expression plasmids, and one day later infected (MOI = 2) with green fluorescent protein (GFP)-encoding VSVΔG backbone virus (purchased from Kerafast). Two hours later, the medium was replaced by medium containing anti-VSV-G antibody (I1-hybridoma, ATCC CRL-2700) to neutralize residual VSV-G input. After 24 h incubation at 32 °C, the supernatants were harvested. To quantify neutralizing antibodies (nAbs), serial dilutions of serum samples were incubated for 1 h at 37 °C with an equal volume of S pseudotyped VSV particles and inoculated on Vero E6 cells (SARS-CoV-2) for 18 h.

The percentage of GFP expressing cells was quantified on a Cell Insight CX5/7 High Content Screening platform (Thermo Fischer Scientific) with Thermo Fisher Scientific HCS Studio (v.6.6.0) software. Neutralization IC50 values were determined by normalizing the serum neutralization dilution curve to a virus (100%) and cell control (0%) and fitting in GraphPad Prism (inhibitor vs. response, variable slope, four parameters model with top and bottom constraints of 100 % and 0 % respectively).

### 2.4 Stimulation of SARS-CoV-2 specific T cells

Heparinized whole blood was used for the EUROIMMUN SARS-CoV-2 interferon gamma release assay (IGRA) kit that was executed according to the manufacturer’s instructions adapted as described below (**Supplementary Figure 1**). Heparinized whole blood was transferred into three different tubes (BLANK, COV2 and STIM) followed by an incubation step for 20-24 h at 37 (±1) °C after inverting 6 times. Following incubation, the tubes were centrifuged at RT for 10 min at 700 g. Approximately 200 µL of stimulated heparinized plasma from each tube was pipetted into Eppendorf tubes and centrifuged again at RT for 10 min at 12,000 g. Finally, the supernatant was pipetted into cryovials and stored at -20 °C until measurement via the EUROIMMUN Quant-T-Cell ELISA (interferon-γ (IFN-γ) ELISA). Importantly, the pellet containing the cellular fraction was used for additional flowcytometry (see 2.5.2 T cell phenotyping and activity).

#### 2.4.1 Interferon γ (IFN-γ) ELISA

Specific SARS-CoV-2 induced IFN-γ release was determined via the EUROIMMUN Quant-T-Cell ELISA according to the manufacturer’s instructions. A photometric measurement was performed at a wavelength of 450 nm with a reference measurement at 620 nm. For each tube, IFN-γ concentrations were determined using a standard curve that was fitted via GraphPad Prism (four parameters model without restrictions). Then, for each subject, the determined IFN-γ concentration from the unstimulated control (BLANK) was subtracted from the determined IFN-γ concentrations of both the stimulation control (STIM) and SARS-CoV-2 stimulated condition (COV2). Lot-specific lyophilized calibrators and controls were used as a standard.

### 2.5 Flowcytometric analyses

#### 2.5.1 Circulating RBD specific B cells

Thawed PBMCs were stained with a selective B cell staining panel that is listed in **Supplementary Table 1**. After a final washing step, the pellets were resuspended in 300 µL dPBS and immediately acquired on the flowcytometer (FACSVerse device, BD Biosciences, USA).

Living B cells were selected from the PBMC pool via a CD3^-^/CD19^+^/Zombie^-^ gating strategy (**Supplementary Figure 2A**). Specific B cell reactivity against SARS-CoV-2 was assessed with wild type RBD-biotin and PE-streptavidin. Within each sample, a negative control tube (without RBD-biotin) was included to correct for sample-specific background. For each staining experiment, a sample with documented RBD specific B cells was used as a positive control for quality assessment.

#### 2.5.2 T cell phenotyping and activity

The remaining cell pellet from the IGRA tubes (see 2.4 Stimulation of SARS-CoV-2 specific T cells) was immediately resuspended with dPBS in a total volume of 500 µL. A whole blood staining was performed on the reconstituted samples. In summary, T cell staining (**Supplementary Table 1**) was performed in 150 µL of the reconstituted samples. Following incubation for 30 min at 4 °C, 3 mL of red blood cell (RBC) lysis buffer (BD FACS^™^ lysing solution, BD Biosciences, USA) was added for 5 min at RT to allow RBC lysis. After extensive washing, the pellets were resuspended in 350 µL dPBS and immediately acquired on the flowcytometer. T cells were selected via gating (**Supplementary Figure 2B**) on the CD3^+^ population and further divided into helper T cells (T_H_; CD4^+^/CD8^-^), circulating follicular helper T cells (T_CFH_; CD4^+^/CD8^-^/CXCLR5^hi^) and cytotoxic T cells (T_C_; CD4^-^/CD8^+^). Membrane markers used to asses T cell activation were CD40L and CD69. The gating strategy was based on the fluorescence minus one (FMO) signal retrieved for each individual fluorochrome. For each subject, besides the condition with SARS-CoV-2 specific antigens, an unstimulated negative (BLANK) and a positive control (STIM) condition were available to respectively correct for background and to assess intrinsic cell functionality.

### 2.6 Viral whole genome sequencing

RNA extraction was performed by using the DEXR-15-LM96 kit for automated extraction (Diagenode, Seraing, Belgium) with 350µL sample input. Extracted RNA was eluted from magnetic beads in 50 μl of UltraPure DNase/RNasefree distilled water. Following RNA extraction, cDNA was synthesized followed by multiplex PCR amplification using a modified version of the ARTIC V3 LoCost protocol with the Midnight primer set (1200 bp amplicons). The libraries were sequenced on a MinION using R9.4.1 flow-cells (Oxford Nanopore Technologies, Oxford, UK) and MinKnow software v21.02.1. The resulting fast5 reads were basecalled and demultiplexed using Guppy v5.0.16 in super accuracy mode. Genome assembly was performed using the ARTIC bioinformatics pipeline v1.1.3, which entails adapter trimming, mapping to the reference strain Wuhan-Hu-1 (MN908947), as previously described (36).

### 2.7 Statistical analyses

Statistical analysis was performed using both Microsoft Excel (version 365 for Windows, Microsoft Corporation, USA) and GraphPad Prism (version 9.0.0 for Windows, GraphPad Software, USA). Flowcytometric data was processed using FCS Express (version 7 research edition for Windows, De Novo Software, USA). Continuous variables were presented as mean ± standard deviation if the Shapiro-Wilk normality test was successful or as either median ± interquartile range (IQR) or median with the interval between quartile 1 and 3 (Q1-Q3) if not. Confidence intervals (CI) of medians were calculated via the standard method described by Zar JH (37). Discrete and categorical variables were shown as respectively numbers or categories with percentages between brackets. Comparison between parameters was done using a student t-test or one-way ANOVA with a post-hoc correction for multiple comparisons via Tukey’s test and after assessment of constant variance using Levene’s test if normality was met or with the respective non-parametric alternatives when not. Paired analyses were done if appropriate. Correlation between parameters was assessed via bivariate analysis expressed via a Pearson determination coefficient (R^2^); a Pearson r or via a Spearman r if there was no normality and also via a multivariate analysis using principal component analysis (PCA). For all tests, the significance was set at a two-tailed probability level of 0.05.

## 3 Results

### 3.1 Trial characteristics and exclusions

All enrolled healthcare workers (N = 32) were Caucasian and exact half of them were women. The age ranged between 25 and 51 years with a mean of 36 ± 7 (95% confidence interval (CI): 33-38) years. During this trial, one subject was excluded for all analyses because she dropped out between t_pre_ and t_3m_ (**Figure 1**). Anti-N IgG titers were measured at both t_pre_ and t_3m_ to exclude (subclinical) natural SARS-CoV-2 infection. For both timepoints, the anti-N IgG titers remained below the index value with a mean of 0.022 ± 0.038 and 0.100 ± 0.113 at respectively t_pre_ and t_3m_.

### 3.2 Antibody and B cell response three months after BNT162b2 vaccination

#### 3.2.1 BNT162b2 vaccination induces functionally active antibodies

Three months after BNT162b2 vaccination, the vaccine-induced antibody response was described via both serology and in vitro neutralization assays. Spike-specific antibodies are represented in **Figure 2**.

**Figure 2.**
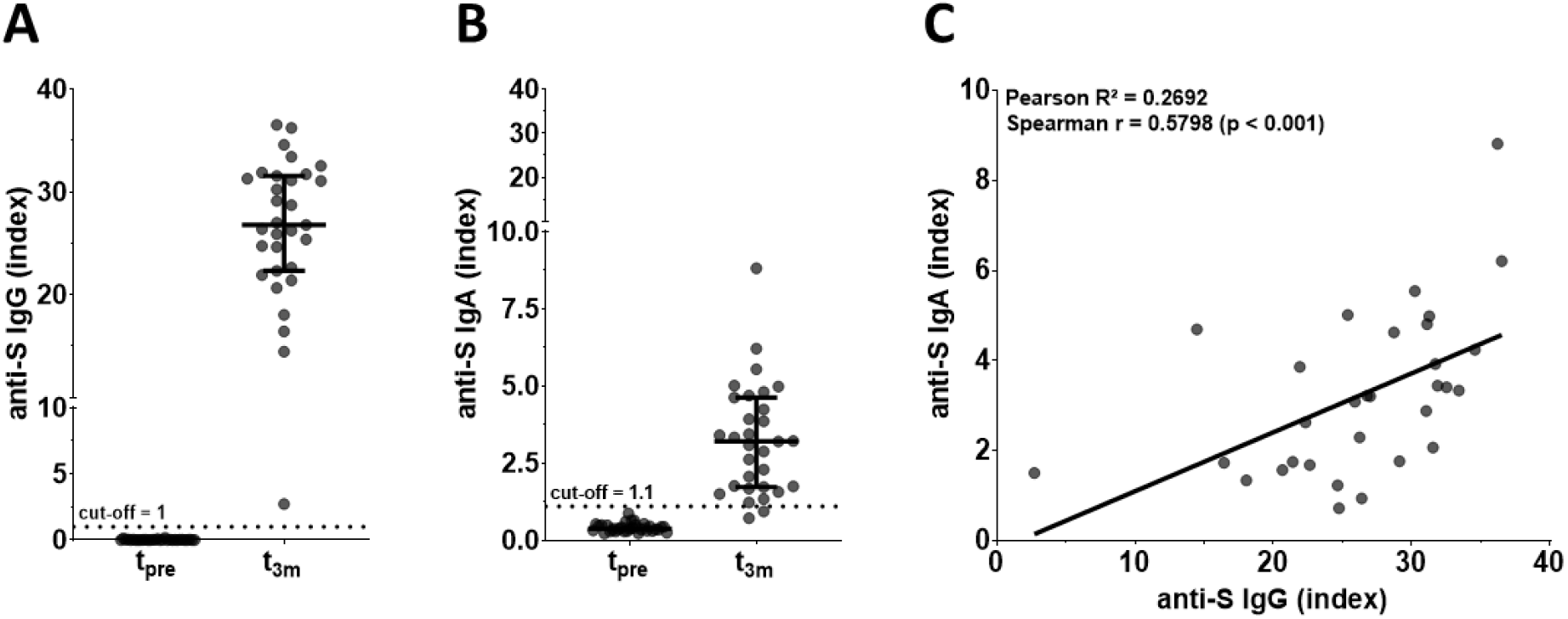
SARS-CoV-2 anti-S serology (N = 31). **(A)** anti-S IgG titers, **(B)** anti-S IgA titers and **(C)** simple linear regression and correlation between anti-S IgG and IgA antibodies 3 months after BNT162b2 vaccination. Error bars represent median with IQR. Abbreviations: S = spike, pre = baseline sampling moment before vaccination, 3m = 3 months after baseline, IQR = interquartile range.

At baseline, all 31 subjects were negative for both anti-S IgG and IgA with a median index titer of 0.010 ± 0.020 (95% CI: 0.000-0.020) and 0.371 ± 0.194 (95% CI: 0.320-0.474) respectively. Three months after vaccination, all individuals showed detectable anti-S IgG antibodies with a median index titer of 26.8 ± 9.23 (95% CI: 24.7-31.1) of which two of these individuals had no anti-S IgA response (**Figure 2A-B**). Anti-S IgG and IgA titers were moderately but significantly correlated with a Spearman r of 0.5798 (p < 0.001) and a Pearson R^2^ of 0.2692 (**Figure 2C**).

Next, anti-RBD IgG titers were determined (**Figure 3A**). In line with the above findings, no anti-RBD IgG antibodies could be detected at baseline (< 160 BAU/mL), whereas all but one had anti-RBD IgG titers three months post-vaccination with a median titer of 827.0 (95% CI: 661.0-1103) with a Q1-Q3 interval of 599.0-1310 BAU/mL.

**Figure 3.**
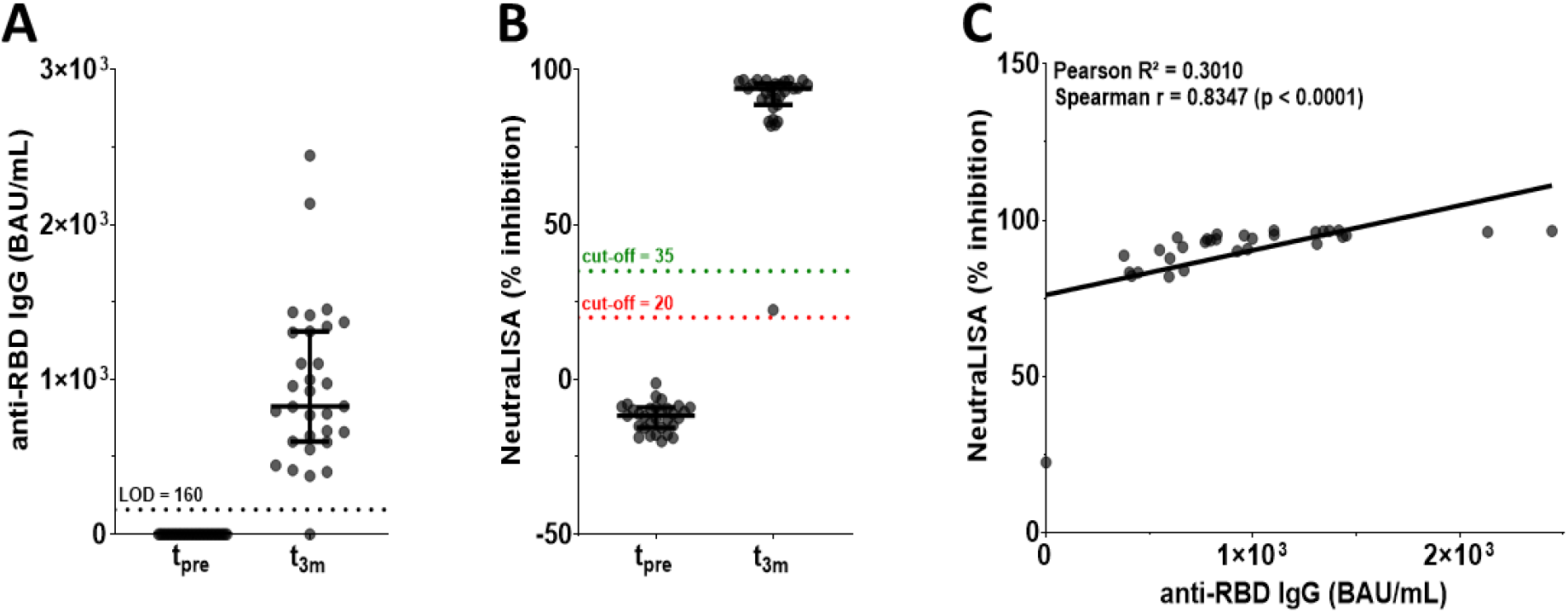
SARS-CoV-2 anti-RBD serology and antibody functionality (N = 31). **(A)** anti-RBD IgG titers, **(B)** in vitro neutralization efficacy and **(C)** simple linear regression and correlation between anti-RBD IgG and antibody functionality at three months post-BNT162b2 vaccination. Error bars represent median with IQR. Abbreviations: RBD = receptor-binding domain, pre = baseline sampling moment before vaccination, 3m = 3 months after baseline, IQR = interquartile range.

Additionally, the capacity of the induced antibodies to neutralize the binding between wild type RBD and human ACE2, was assessed. Overall antibody neutralization capacity was high with a median % inhibition compared to a blank control of 93.91 ± 6.08 (95% CI: 90.45-95.13; **Figure 3B**). A moderate but significant correlation was seen between anti-RBD IgG titers and antibody functionality with a Spearman r of 0.8347 (p < 0.0001) and a Pearson R^2^ of 0.3010 (**Figure 3C**).

#### 3.2.2 Circulating RBD specific B cells

In parallel to the assessment of serology and functionality of the vaccine-induced antibodies, the precursor rate of circulating RBD specific B cells was examined. For each individual, a PBMC aliquot of t_pre_ and t_3m_ were analyzed within the same experiment to allow the detection of vaccine-specific changes. An example of the gating strategy used for the flowcytometric B cell phenotyping is shown in **Supplementary Figure 2A**.

The magnitude of CD3^-^/CD19^+^ B cells was comparable between both timepoints and ranged between 1.93 – 14.16 % of gated lymphocytes. Furthermore, a small (0.087 ± 0.068 % of parent, i.e. living B cells) but significant (p < 0.005) population of circulating RBD specific B cells was detected in 10 of the 31 individuals (32.26 %) at three months post vaccination (**Figure 4A**). Within these 10 subjects, the rate of circulating RBD specific B cells showed a significant good correlation with anti-RBD IgG titers (Pearson R^2^ = 0.6314, Spearman r = 0.8571 with p < 0.005, **Figure 4B**) and a moderate correlation with antibody functionality (Pearson R^2^ = 0.3398, Spearman r = 0.8902 with p < 0.005, **Figure 4C**).

**Figure 4.**
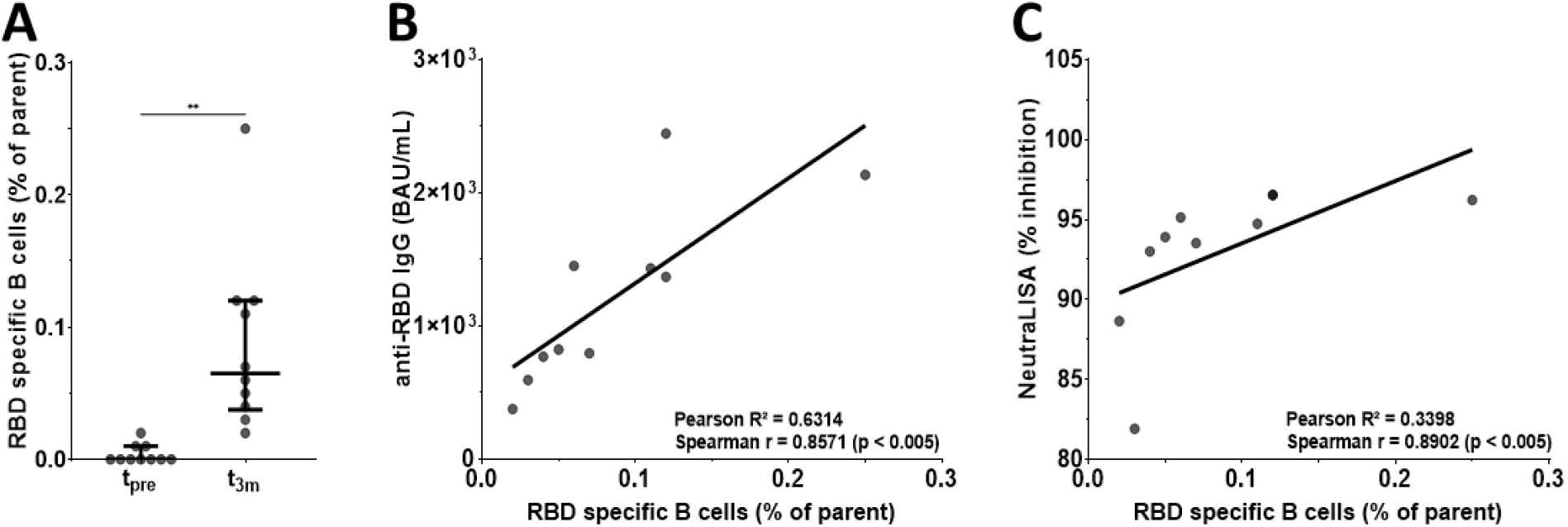
Flowcytometric determination of both B cell phenotype and rate of circulating RBD specific B cells. **(A)** Scatter plot representing the circulating RBD specific B cell populations that were found in 10 participants. Each data point represents the percentage of RBD specific B cells after subtraction of the sample-specific background (i.e. condition with RBD-biotin minus condition without RBD-biotin). ** Wilcoxon test: p < 0.01. **(B)** Simple linear regression and correlation between anti-RBD IgG and circulating RBD specific B cells found in 10 participants at three months post-BNT162b2 vaccination. **(C)** Simple linear regression and correlation between antibody functionality and circulating RBD specific B cells found in 10 participants at three months post-BNT162b2 vaccination. Error bars represent median ± IQR. Abbreviations: RBD = receptor-binding domain, pre = baseline sampling moment before vaccination, 3m = 3 months after baseline, IQR = interquartile range.

### 3.3 T cell immune response three months after BNT162b2 vaccination

#### 3.3.1 BNT162b2 vaccination induces SARS-CoV-2-specific IFN-γ production by T cells

Next, we also defined the SARS-CoV-2 specific T cell response. Firstly, we assessed the vaccine-induced specific IFN-γ release by T cells upon restimulation with SARS-CoV-2 antigens (**Figure 5**). Three subjects were excluded from these analyses due to technical issues.

**Figure 5.**
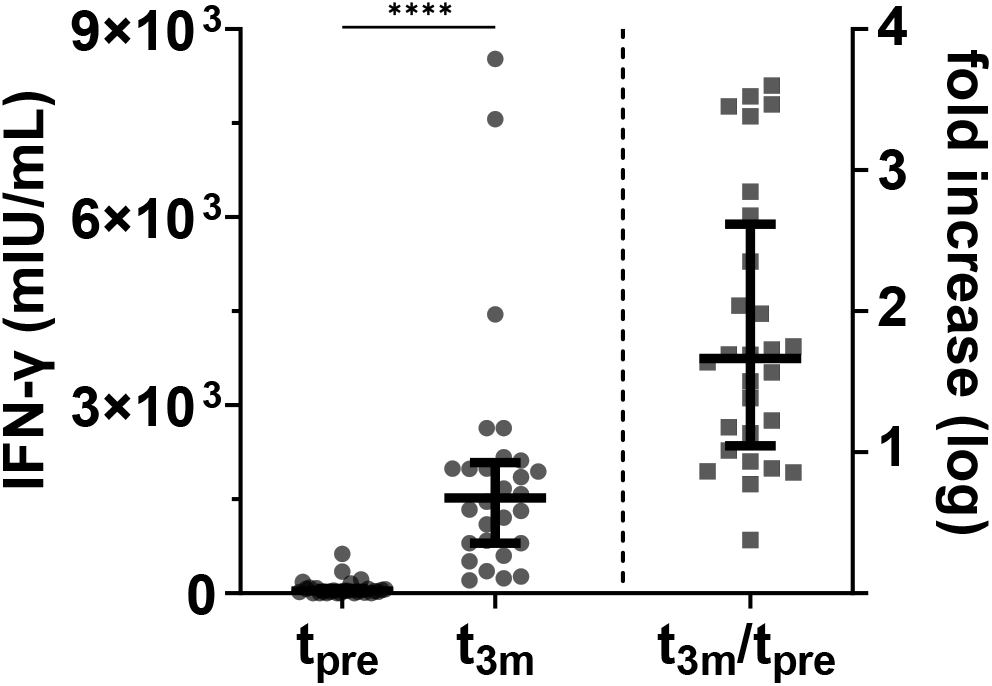
SARS-CoV-2 specific T cell mediated IFN-γ release (N = 28). Scatter plot representing the SARS-CoV-2 specific IFN-γ release after subtraction of the unstimulated negative control at both baseline and 3 months after vaccination. The right part of the plot shows the subject-specific fold increase in SARS-CoV-2 specific IFN-γ release. **** Wilcoxon test: p < 0.0001. Error bars represent median ± IQR. Abbreviations: IFN-γ = interferon-γ, pre = baseline sampling moment before vaccination, 3m = 3 months after baseline, IQR = interquartile range.

The median SARS-CoV-2-specific T cell mediated IFN-γ release post vaccination was 1520 ± 1287 mIU/mL (95% CI: 836.5-1986) with values ranging between 204.8 and 8523 mIU/mL. These IFN-γ values were significantly higher (p < 0.0001) compared to baseline values (median: 34.77 ± 73.50 mIU/mL). Three months post vaccination, there was at least a three-fold increase of SARS-CoV-2-specific T cell mediated IFN-γ release detected and even exceeding 10,000-fold for some participants. At both timepoints, there was a clear IFN-γ release observed in the stimulation control condition (data not shown).

#### 3.3.2 SARS-CoV-2-specific CD4 and CD8 T cells display an activated phenotype after BNT162b2 vaccination

Using the reconstituted cell pellets from the IGRA tubes, we were able to include T cell phenotyping in addition to the SARS-CoV-2 specific IFN-γ release. An example of the applied gating strategy can be retrieved in **Supplementary Figure 2B**. In accordance with the B cell phenotyping, the magnitudes of CD3^+^ lymphocytes (i.e. T cells), CD4^-^/CD8^+^ cytotoxic T cells (T_C_), CD4^+^/CD8^-^ helper T cells (T_H_) and CD4^+^/CD8^-^/CXCR5^hi^ circulating follicular T helper cells (T_CFH_) were similar for all three conditions within the IGRA (data not shown).

Three months after vaccination, a significant upregulation of the early activation marker CD69 was detected upon SARS-CoV-2 specific restimulation in the T_C_ (p < 0.0001), T_H_ (p < 0.0001) and T_CFH_ (p < 0.005) cell subsets (**Figure 6A**). Furthermore, CD40L was also significantly upregulated in both T_H_ (p < 0.0001) and T_CFH_ (p < 0.005) subsets (**Figure 7A**). For each sample, a condition with mitogenic stimulation was analyzed as well as means of quality control (data not shown).

**Figure 6.**
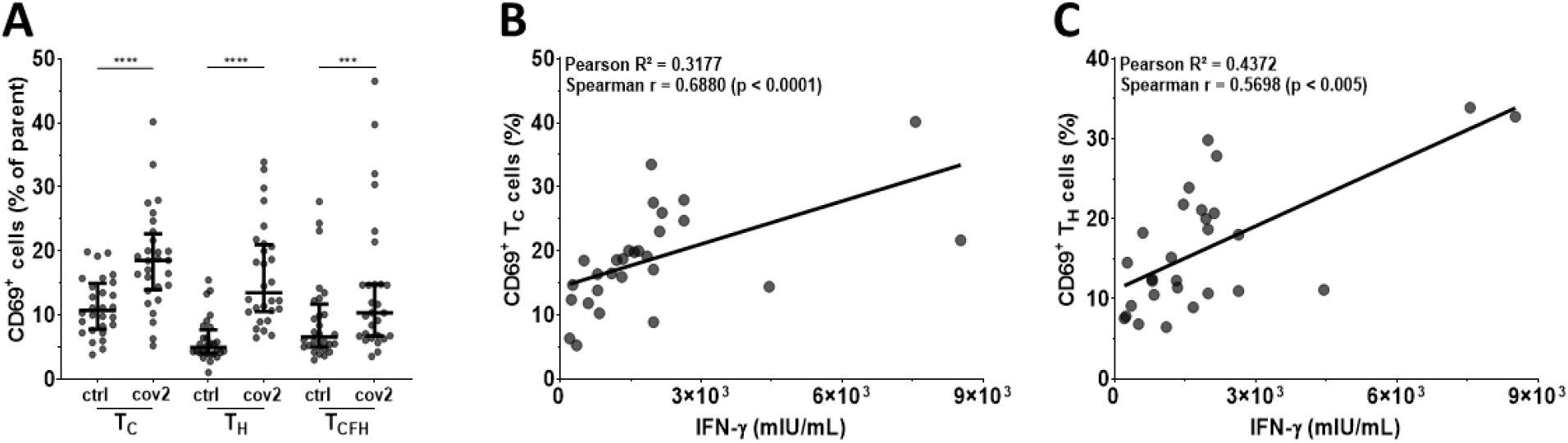
Expression of activation marker CD69 on SARS-CoV-2 specific T cells after BNT162b2 vaccination (N = 28). **(A)** Scatter plot showing the percentages of CD69^+^ T_C_, T_H_ and T_CFH_ cells present in the condition without stimulation (ctrl) and after SARS-CoV-2 specific stimulation (cov2). **** Wilcoxon test: p < 0.0001, *** Wilcoxon test: p < 0.005. **(B-C)** Simple linear regression and correlation between SARS-CoV-2 specific T cell mediated IFN-γ release and following parameters of cellular immunity: **(B)** percentage of CD69^+^ T_C_ cells and **(C)** percentage of CD69^+^ T_H_ cells. Error bars represent median ± IQR. Abbreviations: IFN-γ = interferon-γ, pre = baseline sampling moment before vaccination, 3m = 3 months after baseline, IQR = interquartile range.

**Figure 7.**
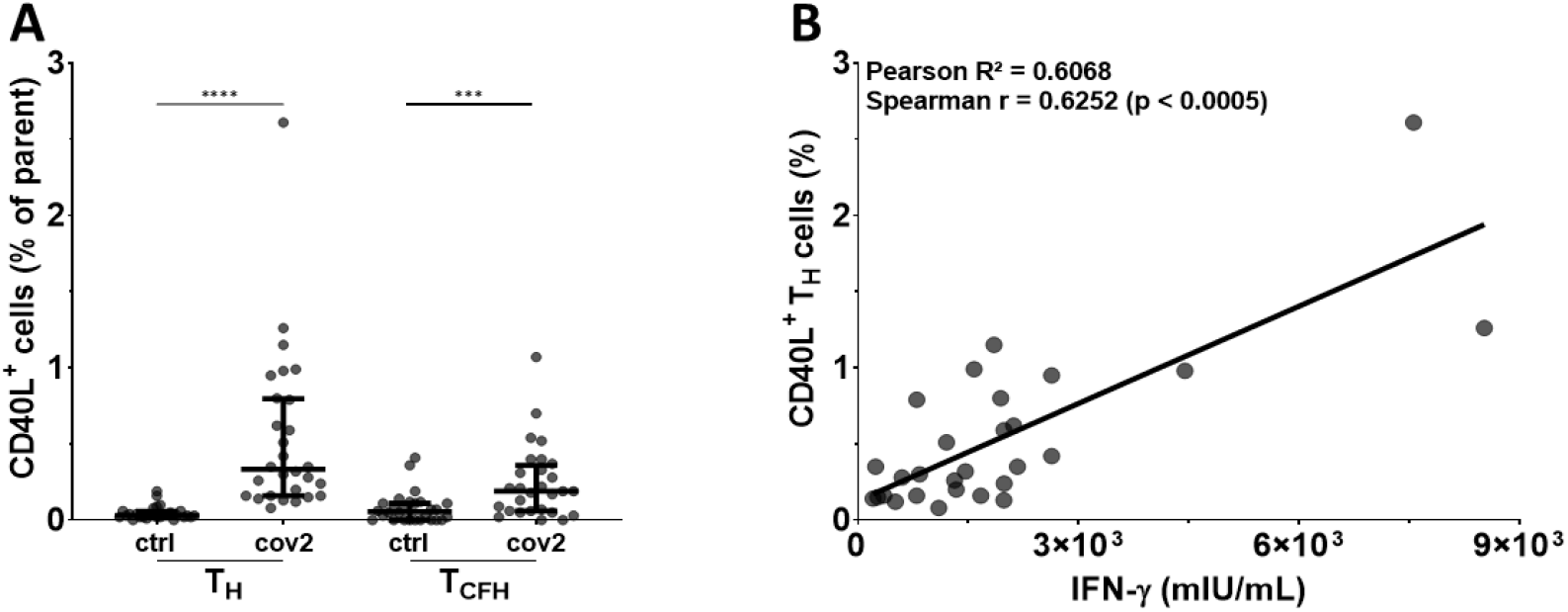
Expression of costimulatory molecule CD40L on SARS-CoV-2 specific T cells after BNT162b2 vaccination (N = 28). **(A)** Scatter plot showing the percentages of CD40L^+^ T_H_ and T_CFH_ cells present in the condition without stimulation (ctrl) and after SARS-CoV-2 specific stimulation (cov2). **** Wilcoxon test: p < 0.0005, *** Wilcoxon test: p < 0.005. **(B)** Simple linear regression and correlation between SARS-CoV-2 specific T cell mediated IFN-γ release and percentage of CD40L^+^ T_H_ cells. Error bars represent median ± IQR. Abbreviations: IFN-γ = interferon-γ, pre = baseline sampling moment before vaccination, 3m = 3 months after baseline, IQR = interquartile range.

SARS-CoV-2 specific T cell mediated IFN-γ release was moderately but significantly correlated with both CD69^+^ on T_C_ (Pearson R^2^ = 0.3177, Spearman r = 0.6880 with p < 0.0001; **Figure 6B**) and T_H_ (Pearson R^2^ = 0.4372, Spearman r = 0.5698 with p < 0.005; **Figure 6C**) and CD40L^+^ expression on T_H_ (Pearson R^2^ = 0.6068, Spearman r = 0.6252 with p < 0.0005; **Figure 7B**).

### 3.4 The magnitudes of B and T cell response are not correlated three months after BNT162b2 vaccination

Finally, we aligned the magnitude of the different SARS-CoV-2 specific B and T cell parameters but no significant linear relations nor correlations could be observed (Pearson R^2^ < 0.15 and Spearman r < 0.40 with p > 0.10 in all cases). Also, within the 10 healthcare workers that had circulating RBD specific B cells, the precursor rate was not correlated either with CD69 activated (Spearman r = 0.0426, p > 0.50) or CD40L activated T cells (Spearman r = 0.1398, p > 0.50). Additionally, PCA was performed to identify parameters that contribute most to the overall variance found within this cohort (**Figure 8**).

**Figure 8.**
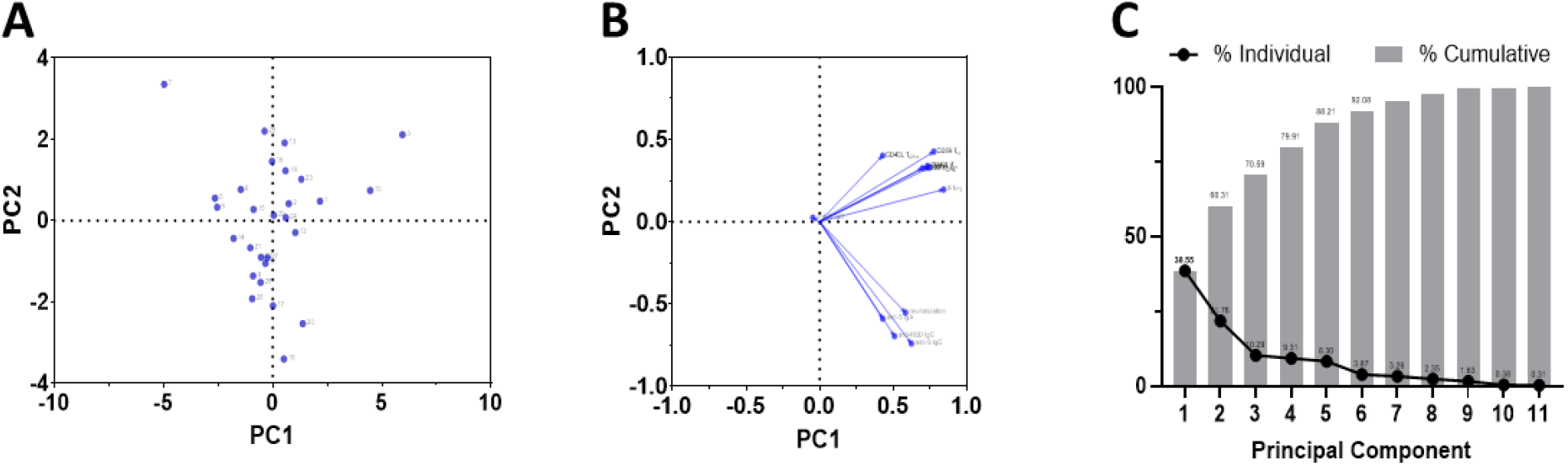
Principal component analysis between parameters of both vaccine-induced humoral and cellular responses (N = 28). **(A)** PC scores plot, **(B)** loadings plot and **(C)** proportions of variance graph. Abbreviations: PCA = principal component analysis, PC = principal component.

First, no clear clustering could be identified from the PC scores plot (**Figure 8A**). Next, PCA revealed that the loadings of all 11 included variables (circulating RBD specific B cells were excluded) appeared on the same side of the loadings plot, showing that each variable correlated positively with PC 1. Also, two different clusters could be identified based on the loadings plot with PC 2 (**Figure 8B**). Combining PC 1 and PC 2 explained approximately 60 % of the overall variance in this cohort (**Figure 8C**).

### 3.5 Symptomatic SARS-CoV-2 BTI despite a vaccine-induced functional antibody response and specific T cell immunity

Within the timeframe of this study, one subject experienced a RT-PCR confirmed symptomatic BTI. This BTI occurred within the collection period for the t_3m_ moment and was therefore redefined as the t_BTI_ timepoint for this participant. As described in **Figure 1**, a nasopharyngeal swab was collected additionally to the blood tubes drawn via venipuncture.

This case concerned a woman in her thirties who presented with general malaise, dry cough and dyspnea 46 days after receiving the booster vaccine. The subject was ill for seven days but without the need for hospitalization. Based on the WHO COVID-19 severity score (38), the subject was classified as experiencing a mild SARS-CoV-2 infection. A few days prior to the BTI, this participant reported to have been in close contact with a hospitalized COVID-19 patient. A complete overview of both the humoral and cellular immune parameters at the t_BTI_ timepoint of this subject are listed in **Table 1**.

**Table 1.**
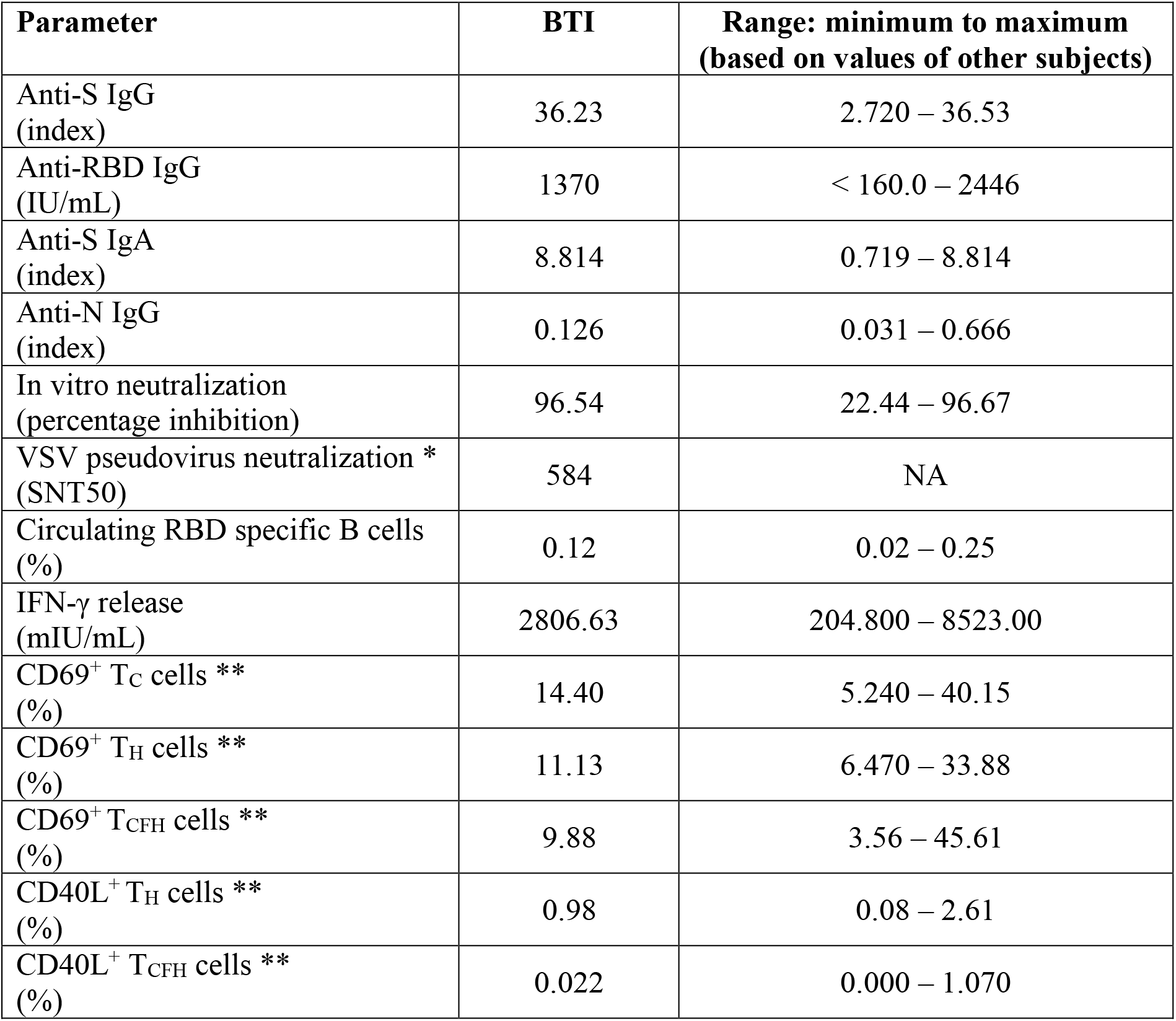
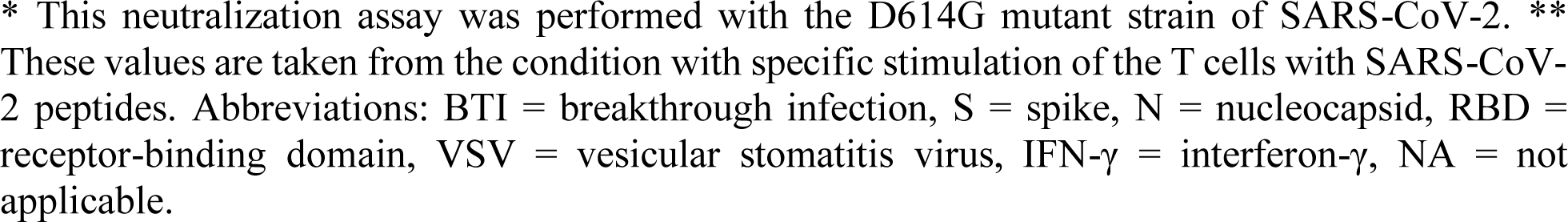
Summary of both humoral and cellular parameters of the subject with symptomatic BTI.

| Parameter | BTI | Range: minimum to maximum<br>(based on values of other subjects) |
| --- | --- | --- |
| Anti-S IgG<br>(index) | 36.23 | 2.720 – 36.53 |
| Anti-RBD IgG<br>(IU/mL) | 1370 | < 160.0 – 2446 |
| Anti-S IgA<br>(index) | 8.814 | 0.719 – 8.814 |
| Anti-N IgG<br>(index) | 0.126 | 0.031 – 0.666 |
| In vitro neutralization<br>(percentage inhibition) | 96.54 | 22.44 – 96.67 |
| VSV pseudovirus neutralization *<br>(SNT50) | 584 | NA |
| Circulating RBD specific B cells<br>(%) | 0.12 | 0.02 – 0.25 |
| IFN-γ release<br>(mIU/mL) | 2806.63 | 204.800 – 8523.00 |
| CD69<sup>+</sup> T<sub>C</sub> cells **<br>(%) | 14.40 | 5.240 – 40.15 |
| CD69<sup>+</sup> T<sub>H</sub> cells **<br>(%) | 11.13 | 6.470 – 33.88 |
| CD69<sup>+</sup> T<sub>CFH</sub> cells **<br>(%) | 9.88 | 3.56 – 45.61 |
| CD40L<sup>+</sup> T<sub>H</sub> cells **<br>(%) | 0.98 | 0.08 – 2.61 |
| CD40L<sup>+</sup> T<sub>CFH</sub> cells **<br>(%) | 0.022 | 0.000 – 1.070 |

In summary, at time of presenting with a symptomatic BTI, this participant displayed a functional serological response (both in vitro neutralization and in pseudovirus assay) as well as SARS-CoV-2 specific T-cell reactivity. Moreover, a population of circulating RBD specific B cells could be observed. In order to investigate whether this BTI could be ascribed to any of the currently known VoC, whole viral genome sequencing was performed on the nasopharyngeal swab. Genome sequencing revealed the presence of the B.1.1.7 Pango lineage, which is also known as the alpha VoC.

## 4 Discussion

In this study, we report on SARS-CoV-2 specific B and T cell immunity in healthcare workers in a supraregional hospital in Belgium three months after BNT162b2 vaccination. Importantly, during this study, both the number of occupied hospital and intensive care beds increased substantially defining the start of the so-called third SARS-CoV-2 infection wave in Belgium (from 15 February 2021 onwards) (39). Besides increasing virus circulation, also the emergence of multiple VoC, including alpha (B.1.1.7), beta (B.1.351) and gamma (P1) variants are known to boost infection rates, in particular since vaccination coverage in the general population was still low at that time (14,40).

Serological findings in this cohort were comparable with those described by other groups that studied the serology status post booster BNT162b2 vaccination at different timepoints, i.e. ranging between 2 and 12 weeks post primary vaccination (41–48). Of note, many of these studies monitored vaccine induced antibody responses in patients suffering from a myriad of pathologies (41–43). In addition, the net effect of vaccination is often difficult to highlight as in many studies subjects with earlier natural SARS-COV-2 infection are not excluded (27,49). Three months post vaccination, spike-specific IgA levels were less pronounced compared to IgG levels, the latter observed in all participants of our cohort. A more rapid waning of SARS-CoV-2 IgA antibodies compared to IgG was reported by Wisnewski et al (50). A significant but moderate correlation between spike-specific IgG and IgA was observed, which was in line with findings by other groups (50–52). However, the key question here is in fact not the net level of antigen specific vaccine-induced antibodies, but whether these antibodies are truly protective and thus able to neutralize docking of SARS-CoV-2 virus particles to the human ACE-2 receptor on target cells. Indeed, nAbs that were able to almost completely block the RBD binding to human ACE-2 could be retrieved in all but one participant, confirming the high efficacy rate of the BNT162b2 vaccine after three months. In line with the findings by others (50,52,53), both anti-S and anti-RBD IgG titers were identified as moderate correlates for antibody functionality. Of note, the borderline in vitro neutralization observed in one subject could not be attributed to assay-specific technical issues, as a similar result was observed in an in-house developed test (data not shown). With both IgG and IgA anti-S antibodies just above detection limit, either the mRNA-vaccine encoded spike protein is less immunogenic in this subject or antibody titers – and most likely also functionality – are decreasing more rapidly. In an environment with high virus circulation, including VoC, this person could be more at risk to develop a BTI. Overall, our findings add to high neutralization efficacy of the BNT162b2 vaccine reported by others at earlier timepoints (51,54). Of note, binding and neutralization assays were performed with the Wuhan variant of SARS-CoV-2 encoded by the BNT162b2 vaccine. In other words, one might show high antibody titers and functional neutralization to the original virus strain that could prove to be biologically less relevant against the mutated target proteins of the emergent VoC.

Currently, there is not yet much information available concerning the dynamics and longevity of humoral immunity upon SARS-CoV-2 vaccination. Circulating antigen specific B cells are considered to be a surrogate marker for the resident antigen specific B cells present within the secondary lymphoid organs that are essential to establish B cell memory (18,55–57). Interestingly and opposite to findings by Ciabattini et al. (18), circulating RBD specific B cells were only detectable in one third of the study participants at 3 months post vaccination. In line with the latter, we were only able to retrieve RBD specific B cells in a fraction of convalescent patients (Imbrechts et al., submitted), again in contrast with findings by others such as the group of Nussenzweig et al (48). Of the 10 individuals that displayed circulating RBD specific B cell clones, a correlation was observed between the abundance of RBD-specific B cells on the one hand and both serology and antibody functionality on the other hand (**Figure 4B-C)**. As no signs of ongoing natural infection were found, these data suggest that at least in these participants, the germinal center driven humoral immunity is still being built up three months post mRNA vaccination. Shifting viewpoint from vaccination to COVID-19 treatment options, our findings suggest that PBMCs from vaccinated donors could serve as a source to sort out RBD specific B cell clones able to produce strong nAbs of therapeutic interest. This technique has already been performed with PBMCs from convalescent individuals and already led to the development of several promising nAbs (e.g. LY-CoV555 (58), REGN10933 and REGN10987 (59), ABBV-2B04 (60), TY027 (61) and 3B8 (Imbrechts et al. submitted)) of which some are being evaluated in phase III trials or are already approved by the FDA.

Specific T cell responses upon BNT162b2 vaccination were already documented in the original phase I/II trial by Sahin et al (24,25). In addition, other groups reported a SARS-CoV-2 specific IFN-γ release in either immunocompromised patients (41,62,63), dialysis patients (45,64) or healthcare workers at different timepoints up to four weeks post vaccination (7,27,28). In line with these results, we could clearly observe SARS-CoV-2 specific IFN-γ release in all subjects up to three months post vaccination. Moreover, we were able to include T cell phenotyping as secondary read-out to the whole blood IGRA, hereby eliminating the need for PBMCs nor expensive peptide pools covering the most prominent MHC haplotypes in a given population. Using this approach, we were able to show SARS-CoV-2 specific activation of both T_C_ and T_H_ cell subsets which implies that the SARS-CoV-2 specific antigens in this assay were presented on both MHC class I and II proteins respectively. This was also shown in reports from both Braun et al. (65) and Giminez et al. (66) in which they used a related SARS-CoV-2 antigen pool to stimulate T cells and found respectively an upregulation of activated CD4^+^ and CD8^+^ T cells. Besides membrane expression of CD69 as general early T cell activation marker, we were able to show upregulation of CD40L both in total T_H_ and in the CXCR5^+^ T_CFH_ subsets. Both early T cell activation and membrane CD40L expression are correlated with the release of type II interferon, confirming that BNT162b2 vaccination induces a T_H1_ response (25). In addition, as CD40L is one of the key costimulatory molecules involved in both antibody class switching and BCR affinity maturation (55), CD40L expression on T_H_ and in particular on CXCR5+ T_CFH_ (47) suggests that in vivo antigen re-exposure will most likely lead to a massive and rapid T cell dependent antibody recall response, the latter being exactly the overall goal of vaccination.

Next, we addressed whether the humoral and cellular immune parameters measured in this study were associated to each other. From the PCA analysis, two different clusters could be identified based on the loadings plot with PC 2. Interestingly, these clusters included all the parameters determined within B or T cell immunity. This suggests that measuring only one or two parameters from both the humoral and cellular immune response is sufficient to respectively predict 60 to 80 percent of the overall variability observed in this cohort (**Figure 8C**). Also, the approximate angle of 90 degrees between both clusters emphasizes the absence of any correlation between each other.

Within the three month monitoring time window in this cohort, there was one subject who developed a mild symptomatic BTI (incidence of 3.22 %) although the incidence rate reported in the BNT162b2 phase III trial was very low (< 0.05 %, 8 cases out of 18,198) (67). Of note, one must be careful not to overinterpret this high incidence rate due to the small size of this study. In addition, healthcare workers might be more at risk than the general population, especially during a new infection wave including emergent VoC. Although, it must be mentioned that a recently published report of the Belgian Healthcare Knowledge Center – that included surveillance data since the start of the vaccination campaign in Belgium – mentioned that the national incidence of symptomatic BTI within the general population (of which 71.2% were vaccinated with Comirnaty^®^) was only 0.20 % (68). Antonelli et al. stated that vaccinated people who developed a BTI had mostly milder symptoms and were approximately half less likely to report symptoms of Long COVID-19 Syndrome than infected unvaccinated people (69). This was also the case in our subject as she had a mild infection and recovered completely after seven days. Interestingly, this subject showed a clear vaccine-induced immune response including SARS-CoV-2 specific T cell activity, positive titers of both anti-S and anti-RBD IgG antibodies with a high neutralization efficacy (confirmed in an established pseudovirus assay) and had a small but clearly detectable population of circulating RBD specific B cells. Here, the latter could very well represent the recall response to natural infection in this subject. Whole genome sequencing retrieved that the subject was infected with the alpha variant which was at that time the upcoming VoC within the first phase of the third wave in Belgium. Antibody repertoire profiling of the subject with BTI compared to a pool of age and gender matched controls could yield interesting information, in particular when compared to other vaccine BTI profiles.

At last, it must be noted that this trial has several limitations. Firstly, the number of subjects is rather limited and was based on a power analysis for the serological and IFN-γ release read-outs. Hence, this design is less suited to pick up rather unexpected events such as BTI. Larger studies that look at both arms of the adaptive immune response months after vaccination are needed to further finetune our findings and to characterize BTI more reliable. Secondly, only SARS-CoV-2 naive individuals were included in this study. The pronounced recall response due to vaccination in convalescent individuals has been described and even led to the discussion whether these individuals should receive one rather than two vaccine doses. Addressing this was beyond our scope as we specifically aimed to study the BNT162b2 vaccine-induced de novo immune response in a real-world setting. Thirdly, the emergence of new and potentially more dangerous SARS-CoV-2 variants leads the binding and neutralization assays available at any given time. In this perspective, running neutralization assays with VoC RBD could give deeper insights in the breadth of the vaccine-induced functional immunity. Finally, long-term sustainability of the vaccine-induced immune response can only be truly considered when looking even beyond the three month timepoint. This will be particularly relevant as we are slowly progressing from a global pandemic state to a genuine endemic circulation of the SARS-CoV-2 virus.

## 5 Conclusion

Three months post BNT162b2 mRNA vaccination, previously naive healthcare workers show functional but individually distinct humoral and cellular immune responses to SARS-CoV-2 that do not guarantee protection against the emerging VoC.

## Data Availability

All data produced in the present study are available upon reasonable request to the authors

https://www.gisaid.org

## 6 Conflict of Interest

The authors ZD and KD declare a possible conflict of interest as they are employees of the Institut für Experimentelle Immunologie affiliated to EUROIMMUN Medizinische Labordiagnostika AG (Lübeck, Germany). This company provided following materials for this study: EUROIMMUN SARS-CoV-2 IGRA kit (no. ET 2606-3003), EUROIMMUN interferon-gamma ELISA (no. EQ 6841-9601), EUROIMMUN SARS-CoV-2 NeutraLISA (no. EI 2606-9601-4) and in-house produced RBD-biotin.

## 7 Author Contributions

BC, NC, MI, NG and WM contributed to the conception and design of the manuscript. BC, KC, AV, NC, JVC, XB, MI, NG and WM contributed to the acquisition, analysis or interpretation of data. BC, KC, AV, NC, MI, ZD, KD, KV, NG, SDM and WM have drafted the work or substantively revised it.

## 8 Funding

Internal KU Leuven funding

## 9 Acknowledgments

The authors would like to thank the people from BIOGNOST CV, Heule (Belgium) for their support.

## Notes

### Competing Interest Statement

The authors ZD and KD declare a possible conflict of interest as they are employees of the Institut fur Experimentelle Immunologie affiliated to EUROIMMUN Medizinische Labordiagnostika AG (Lubeck, Germany). This company provided following materials for this study: EUROIMMUN SARS-CoV-2 IGRA kit (no. ET 2606-3003), EUROIMMUN interferon-gamma ELISA (no. EQ 6841-9601), EUROIMMUN SARS-CoV-2 NeutraLISA (no. EI 2606-9601-4) and in-house produced RBD-biotin.

### Clinical Trial

This clinical trial was registered at the EU Clinical Trial Register with ID 2021-001304-15.

### Funding Statement

This study was funded by KU Leuven

## References

1. WHO. World Health Organization: Coronavirus [Internet]. 2021. Available from: https://www.who.int/health-topics/coronavirus#tab=tab_1

2. Corman VM, Landt O, Kaiser M, Molenkamp R, Meijer A, Chu DKW, et al. Detection of 2019 novel coronavirus (2019-nCoV) by real-time RT-PCR. Eurosurveillance. 2020 Jan 23;25(3):2000045.

3. Zhang Y-Z. Novel 2019 coronavirus genome -SARS-CoV-2 coronavirus - Virological [Internet]. Virological.org. 2020. p. 1–7. Available from: https://virological.org/t/novel-2019-coronavirus-genome/319

4. OMS. IHR Emergency Committee on Novel Coronavirus (2019-nCoV) [Internet]. Geneva, Switzerland. 2020. p. 1–4. Available from: https://www.who.int/director-general/speeches/detail/who-director-general-s-statement-on-ihr-emergency-committee-on-novel-coronavirus-(2019-ncov)

5. Romani G, Dal Mas F, Massaro M, Cobianchi L, Modenese M, Barcellini A, et al. Population Health Strategies to Support Hospital and Intensive Care Unit Resiliency during the COVID-19 Pandemic: The Italian Experience. Popul Health Manag. 2021 Apr 1;24(2):174–81.

6. FDA. Pfizer-BioNTech COVID-19 Vaccine Public Assessment Report. 2021.

7. EMA - Committee for Medicinal Products for Human Use. Pfizer-BioNTech COVID-19 Vaccine EMA Public Assesment Report. 2021.

8. EMA - Committee for Medicinal Products for Human Use. Moderna COVID-19 Vaccine EMA Public Assessment Report. 2021.

9. FDA. Moderna COVID-19 Vaccine Public Assessment Report. 2021.

10. EMA - Committee for Medicinal Products for Human Use. AstraZeneca EMA Public Assessment Report. Vol. 31. 2021.

11. EMA - Committee for Medicinal Products for Human. COVID-19 Vaccine Janssen EMA Public Assessment Report. Vol. 31. 2021.

12. FDA. Janssen COVID-19 Vaccine Public Assesment Report. 2021.

13. Study of mRNA Vaccine Formulation Against COVID-19 in Healthy Adults 18 Years of Age and Older - Full Text View - ClinicalTrials.gov [Internet]. 2021. Available from: https://clinicaltrials.gov/ct2/show/NCT04904549?term=sanofi+gsk&cond=SARS-CoV-2&draw=2&rank=1

14. WHO. Tracking SARS-CoV-2 Variants [Internet]. WHO. 2021. p. 1–13. Available from: https://www.who.int/en/activities/tracking-SARS-CoV-2-variants/

15. Otto SP, Day T, Arino J, Colijn C, Dushoff J, Li M, et al. The origins and potential future of SARS-CoV-2 variants of concern in the evolving COVID-19 pandemic. Curr Biol. 2021 Jul 26;31(14):R918–29.

16. Kim Y, Gaudreault NN, Meekins DA, Perera KD, Bold D, Trujillo JD, et al. Effects of Spike Mutations in SARS-CoV-2 Variants of Concern on Human or Animal ACE2-Mediated Virus Entry and Neutralization. bioRxiv Prepr Serv Biol [Internet]. 2021 Aug 25; Available from: https://pubmed.ncbi.nlm.nih.gov/34462749/

17. Bettini E, Locci M. SARS-CoV-2 mRNA Vaccines: Immunological mechanism and beyond. Vaccines. 2021 Feb 1;9(2):1–20.

18. Ciabattini A, Pastore G, Fiorino F, Polvere J, Lucchesi S, Pettini E, et al. Evidence of SARS-CoV-2-Specific Memory B Cells Six Months After Vaccination With the BNT162b2 mRNA Vaccine. Front Immunol. 2021 Sep 28;12:3751.

19. Forgacs D, Jang H, Abreu RB, Hanley HB, Gattiker JL, Jefferson AM, et al. SARS-CoV-2 mRNA Vaccines Elicit Different Responses in Immunologically Naïve and Pre-Immune Humans. Front Immunol. 2021 Sep 27;12:4000.

20. Hammarlund E, Lewis MW, Hansen SG, Strelow LI, Nelson JA, Sexton GJ, et al. Duration of antiviral immunity after smallpox vaccination. Nat Med. 2003 Sep 1;9(9):1131–7.

21. Kedzierska K, Koutsakos M. The ABC of major histocompatibility complexes and T cell receptors in health and disease. Viral Immunol. 2020 Apr 1;33(3):160–78.

22. Gil-Manso S, Carbonell D, López-Fernández L, Miguens I, Alonso R, Buño I, et al. Induction of High Levels of Specific Humoral and Cellular Responses to SARS-CoV-2 After the Administration of Covid-19 mRNA Vaccines Requires Several Days. Front Immunol. 2021 Oct 4;12:3970.

23. Lombardi A, Bozzi G, Ungaro R, Villa S, Castelli V, Mangioni D, et al. Mini Review Immunxsological Consequences of Immunization With COVID-19 mRNA Vaccines: Preliminary Result. Vol. 12, Frontiers in Immunology. Frontiers Media S.A.; 2021. p. 677.

24. Mulligan MJ, Lyke KE, Kitchin N, Absalon J, Gurtman A, Lockhart S, et al. Phase I/II study of COVID-19 RNA vaccine BNT162b1 in adults. Nature. 2020 Aug 12;586(7830):589–93.

25. Sahin U, Muik A, Derhovanessian E, Vogler I, Kranz LM, Vormehr M, et al. COVID-19 vaccine BNT162b1 elicits human antibody and TH1 T cell responses. Nature. 2020 Sep 30;586(7830):594–9.

26. Vogel AB, Kanevsky I, Che Y, Swanson KA, Muik A, Vormehr M, et al. BNT162b vaccines protect rhesus macaques from SARS-CoV-2. Nature. 2021 Apr 8;592(7853):283– 9.

27. Casado JL, Haemmerle J, Vizcarra P, Rodriguez-Dominguez M, Velasco T, Velasco H, et al. T-cell response after first dose of BNT162b2 SARS-CoV-2 vaccine among healthcare workers with previous infection or cross-reactive immunity. Clin Transl Immunol. 2021;10(9).

28. Painter MM, Mathew D, Goel RR, Apostolidis SA, Pattekar A, Kuthuru O, et al. Rapid induction of antigen-specific CD4+ T cells is associated with coordinated humoral and cellular immunity to SARS-CoV-2 mRNA vaccination. Immunity. 2021 Sep 14;54(9):2133-2142.e3.

29. Glatman-Freedman A, Bromberg M, Dichtiar R, Hershkovitz Y, Keinan-Boker L. The BNT162b2 vaccine effectiveness against new COVID-19 cases and complications of breakthrough cases: A nation-wide retrospective longitudinal multiple cohort analysis using individualised data. EBioMedicine. 2021 Oct 1;72.

30. Blanquart F, Abad C, Ambroise J, Bernard M, Cosentino G, Giannoli JM, et al. Characterisation of vaccine breakthrough infections of sars-cov-2 delta and alpha variants and within-host viral load dynamics in the community, france, june to july 2021. Eurosurveillance. 2021 Sep 16;26(37).

31. Tene Y, Levytskyi K, Adler A, Halutz O, Paran Y, Goldshmidt H, et al. An outbreak of SARS-CoV-2 infections among hospital personnel with high mRNA vaccine uptake. Infect Control Hosp Epidemiol. 2021

32. Bahl A, Johnson S, Maine G, Garcia MH, Nimmagadda S, Qu L, et al. Vaccination reduces need for emergency care in breakthrough COVID-19 infections: A multicenter cohort study. Lancet Reg Heal - Am. 2021 Sep;100065.

33. EMA. Committee for Medicinal Products for Human Use. Guideline on bioanalytical method validation. EMEA/CHMP/EWP/192217/2009 Rev 1 Corr 2** [Internet]. 2012;1–23. Available from: www.ema.europa.eu/contact

34. NIBSC NI for BS andControls. WHO International Standard First WHO International Standard for anti-SARS-CoV-2 immunoglobulin (human) NIBSC code: 20/136 Instructions for use (Version 2.0, Dated 17/12/2020) [Internet]. 2020. Available from: http://www.nibsc.org/standardisation/international_standards.aspx

35. Sanchez-Felipe L, Vercruysse T, Sharma S, Ma J, Lemmens V, Van Looveren D, et al. A single-dose live-attenuated YF17D-vectored SARS-CoV-2 vaccine candidate. Nature. 2021 Feb 11;590(7845):320–5.

36. Van Cleemput J, van Snippenberg W, Lambrechts L, Dendooven A, D’Onofrio V, Couck L, et al. Organ-specific genome diversity of replication-competent SARS-CoV-2. Nat Commun. 2021 Nov 16;12(1):1–11.

37. Zar JH. Biostatistical Analysis. 5th ed. 2010.

38. Diaz, Janet; Appiah, John; Askie, Lisa; Baller, April; Banerjee, Anshu; Barkley, Shannon; Bertagnolio, Silvia; Hemmingsen, Bianca; Bonet, Mercedes; Cunningham J. COVID-19 : Clinical management Living guidance [Internet]. World Health Organization. 2021. p. 81. Available from: https://www.who.int/publications/i/item/WHO-2019-nCoV-clinical-2021-1

39. Sciensano. Covid-19 Surveillance Frequently Asked Questions [Internet]. 2021. Available from: https://covid-19.sciensano.be/sites/default/files/Covid19/COVID-19_FAQ_ENG_final.pdf

40. Baele G, Cuypers L, Maes P, Dellicour S, Keyaerts E, Wollants E, et al. Genomic surveillance of SARS-CoV-2 in Belgium [Internet]. Vol. 2021. 2021. Available from: https://www.uzleuven.be/nl/laboratoriumgeneeskunde/genomic-surveillance-sars-cov-2-belgium

41. Amodio D, Ruggiero A, Sgrulletti M, Pighi C, Cotugno N, Medri C, et al. Humoral and Cellular Response Following Vaccination With the BNT162b2 mRNA COVID-19 Vaccine in Patients Affected by Primary Immunodeficiencies. Front Immunol. 2021 Oct 4;12:3947.

42. Braun-Moscovici Y, Kaplan M, Braun M, Markovits D, Giryes S, Toledano K, et al. Disease activity and humoral response in patients with inflammatory rheumatic diseases after two doses of the Pfizer mRNA vaccine against SARS-CoV-2. Ann Rheum Dis. 2021 Oct 1;80(10):1317–21.

43. Achiron A, Mandel M, Dreyer-Alster S, Harari G, Magalashvili D, Sonis P, et al. Humoral immune response to COVID-19 mRNA vaccine in patients with multiple sclerosis treated with high-efficacy disease-modifying therapies. Ther Adv Neurol Disord. 2021;14.

44. Grupper A, Sharon N, Finn T, Cohen R, Israel M, Agbaria A, et al. Humoral response to the pfizer bnt162b2 vaccine in patients undergoing maintenance hemodialysis. Clin J Am Soc Nephrol. 2021 Jul 1;16(7):1037–42.

45. Zitt E, Davidovic T, Schimpf J, Abbassi-Nik A, Mutschlechner B, Ulmer H, et al. The Safety and Immunogenicity of the mRNA-BNT162b2 SARS-CoV-2 Vaccine in Hemodialysis Patients. Front Immunol. 2021 Jun 16;12:2390.

46. Lustig Y, Sapir E, Regev-Yochay G, Cohen C, Fluss R, Olmer L, et al. BNT162b2 COVID-19 vaccine and correlates of humoral immune responses and dynamics: a prospective, single-centre, longitudinal cohort study in health-care workers. Lancet Respir Med. 2021 Sep 1;9(9):999–1009.

47. Segundo DS, Comins-Boo A, Irure-Ventura J, Renuncio-García M, Roa-Bautista A, González-Lípez E, et al. Immune assessment of BNT162b2 m-RNA-spike based vaccine response in adults. Biomedicines. 2021 Aug 1;9(8).

48. Wang Z, Schmidt F, Weisblum Y, Muecksch F, Barnes CO, Finkin S, et al. mRNA vaccine-elicited antibodies to SARS-CoV-2 and circulating variants. Nature. 2021 Apr 22;592(7855):616–22.

49. Hirotsu Y, Amemiya K, Sugiura H, Shinohara M, Takatori M, Mochizuki H, et al. Robust Antibody Responses to the BNT162b2 mRNA Vaccine Occur Within a Week After the First Dose in Previously Infected Individuals and After the Second Dose in Uninfected Individuals. Front Immunol. 2021 Aug 26;12:3457.

50. Wisnewski A V., Luna JC, Redlich CA. Human IgG and IgA responses to COVID-19 mRNA vaccines. PLoS One. 2021 Jun 1;16(6 June):e0249499.

51. Pratesi F, Caruso T, Testa D, Tarpanelli T, Gentili A, Gioè D, et al. Bnt162b2 mrna sars-cov-2 vaccine elicits high avidity and neutralizing antibodies in healthcare workers. Vaccines. 2021 Jun 1;9(6).

52. Salvagno GL, Henry BM, Di Piazza G, Pighi L, De Nitto S, Bragantini D, et al. Anti-spike s1 iga, anti-spike trimeric igg, and anti-spike rbd igg response after bnt162b2 covid-19 mrna vaccination in healthcare workers. J Med Biochem. 2021;40(4):327–34.

53. Naaber P, Tserel L, Kangro K, Sepp E, Jürjenson V, Adamson A, et al. Dynamics of antibody response to BNT162b2 vaccine after six months: a longitudinal prospective study. Lancet Reg Heal - Eur. 2021 Sep;100208.

54. Favresse J, Gillot C, Di Chiaro L, Eucher C, Elsen M, Van Eeckhoudt S, et al. Neutralizing antibodies in covid-19 patients and vaccine recipients after two doses of bnt162b2. Viruses. 2021 Jul 1;13(7).

55. Byazrova M, Yusubalieva G, Spiridonova A, Efimov G, Mazurov D, Baranov K, et al. Pattern of circulating SARS-CoV-2-specific antibody-secreting and memory B-cell generation in patients with acute COVID-19. Clin Transl Immunol. 2021 Jan 1;10(2):e1245

56. Turner JS, O’Halloran JA, Kalaidina E, Kim W, Schmitz AJ, Zhou JQ, et al. SARS-CoV-2 mRNA vaccines induce persistent human germinal centre responses. Nature. 2021 Jun 28;596(7870):109–13.

57. Dan JM, Mateus J, Kato Y, Hastie KM, Yu ED, Faliti CE, et al. Immunological memory to SARS-CoV-2 assessed for up to 8 months after infection. Science. 2021 Feb 5;371(6529).

58. An EUA for Bamlanivimab - A Monoclonal Antibody for COVID-19. Vol. 325, JAMA -Journal of the American Medical Association. JAMA; 2021. p. 880–1.

59. An EUA for Casirivimab and Imdevimab for COVID-19 | The Medical Letter, Inc. [Internet]. 2021. Available from: https://secure.medicalletter.org/w1614a

60. Alsoussi WB, Turner JS, Case JB, Zhao H, Schmitz AJ, Zhou JQ, et al. A Potently Neutralizing Antibody Protects Mice against SARS-CoV-2 Infection. J Immunol. 2020 Aug 15;205(4):915–22.

61. NCT04649515. Efficacy and Safety of TY027, a Treatment for COVID-19, in Humans. https://clinicaltrials.gov/show/NCT04649515 [Internet]. 2020; Available from: https://clinicaltrials.gov/ct2/show/NCT04649515

62. Moyon Q, Sterlin D, Miyara M, Anna F, Mathian A, Lhote R, et al. BNT162b2 vaccine-induced humoral and cellular responses against SARS-CoV-2 variants in systemic lupus erythematosus. Ann Rheum Dis. 2021 Oct 4;annrheumdis-2021-221097.

63. Lasagna A, Agustoni F, Percivalle E, Borgetto S, Paulet A, Comolli G, et al. A snapshot of the immunogenicity, efficacy and safety of a full course of BNT162b2 anti-SARS-CoV-2 vaccine in cancer patients treated with PD-1/PD-L1 inhibitors: a longitudinal cohort study. ESMO Open. 2021 Oct;6(5):100272.

64. Van Praet J, Reynders M, De Bacquer D, Viaene L, Schoutteten M, Caluwé R, et al. Predictors and Dynamics of the Humoral and Cellular Immune Response to SARS-CoV-2 mRNA Vaccines in Hemodialysis Patients: A Multicenter Observational Study. J Am Soc Nephrol. 2021 Sep 29;ASN.2021070908.

65. Braun J, Loyal L, Frentsch M, Wendisch D, Georg P, Kurth F, et al. SARS-CoV-2-reactive T cells in healthy donors and patients with COVID-19. Nature. 2020 Jul 29;587(7833):270–4.

66. Giménez E, Albert E, Torres I, Remigia MJ, Alcaraz MJ, Galindo MJ, et al. SARS-CoV-2-reactive interferon-γ-producing CD8+ T cells in patients hospitalized with coronavirus disease 2019. J Med Virol. 2021 Jan 1;93(1):375–82. /

67. European Medicines Agency. Comirnaty | European Medicines Agency [Internet]. Ema. 2021. Available from: https://www.ema.europa.eu/en/medicines/human/EPAR/comirnaty

68. Jespers V, Leroy R, Hulstaert F, Wyndham Thomas C, Van Montfort T, Van Damme UAntwerpen P, et al. Rapid Review of the Evidence on a COVID-19 Booster Dose After a Primary Vaccination Scheule Report for the Task Force Vaccination (Version With Summary in Dutch). 2021.

69. Antonelli M, Penfold RS, Merino J, Sudre CH, Molteni E, Berry S, et al. Risk factors and disease profile of post-vaccination SARS-CoV-2 infection in UK users of the COVID Symptom Study app: a prospective, community-based, nested, case-control study. Lancet Infect Dis. 2021 Sep

