## Supplementary Figures and Tables for "Real-world monitoring of BNT162b2 vaccine-induced SARS-CoV-2 B and T cell immunity in naive healthcare workers: a prospective single center study"

### *Supplementary Material*

#### 1 Supplementary Figures and Tables

##### 1.1 Supplementary Figures

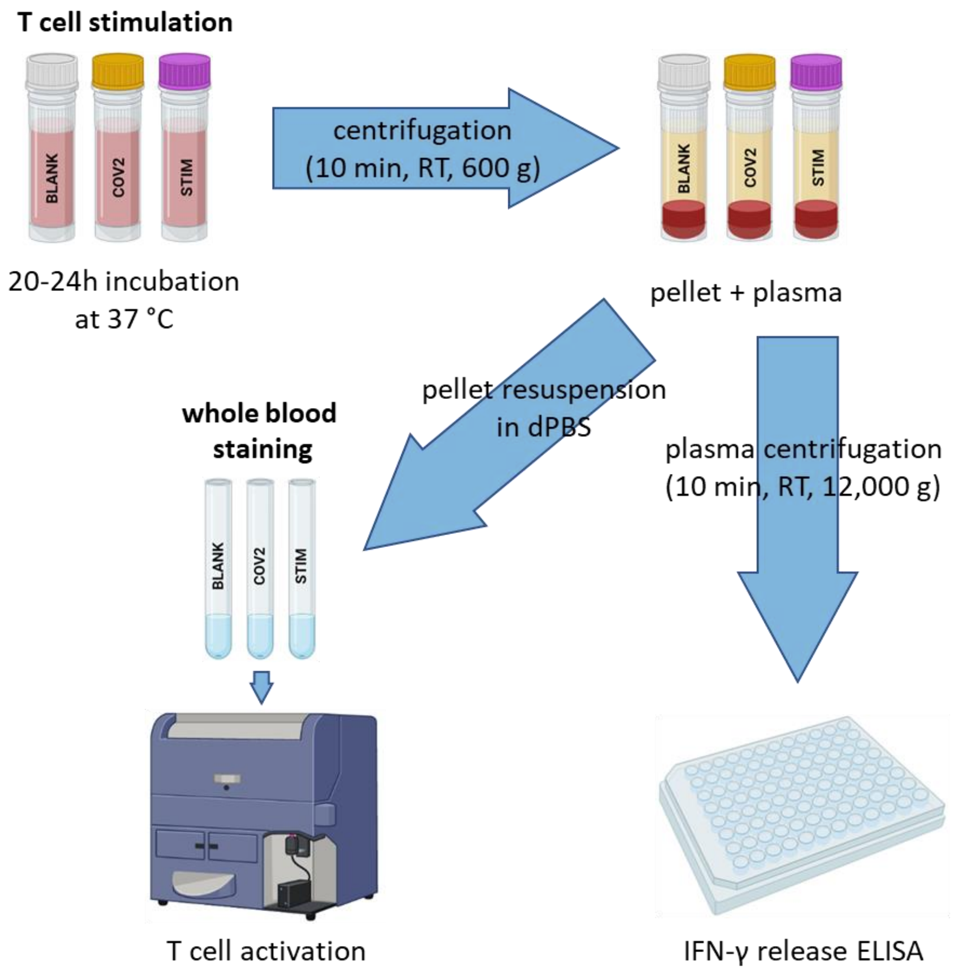


**Supplementary Figure 1. Graphic representation of the secondary read-out procedure to determine SARS-CoV-2 specific T cell activity.** Whereas the plasma was used to determine SARS-CoV-2-specific IFN-γ release via the commercial Quant-T-Cell ELISA (EUROIMMUN, no. EQ 6841-9601), the remaining pellets within the IGRA tubes (BLANK, COV2, STIM) of the SARS-CoV-2 IGRA set (EUROIMMUN, no. ET 2606-3003) were resuspended and stained via a whole-blood principle to assess SARS-CoV-2-specific T cell activity. Abbreviations: RT = room temperature, IFN-γ = interferon γ, ELISA = enzyme-linked immunosorbent assay, IGRA = interferon γ release assay.

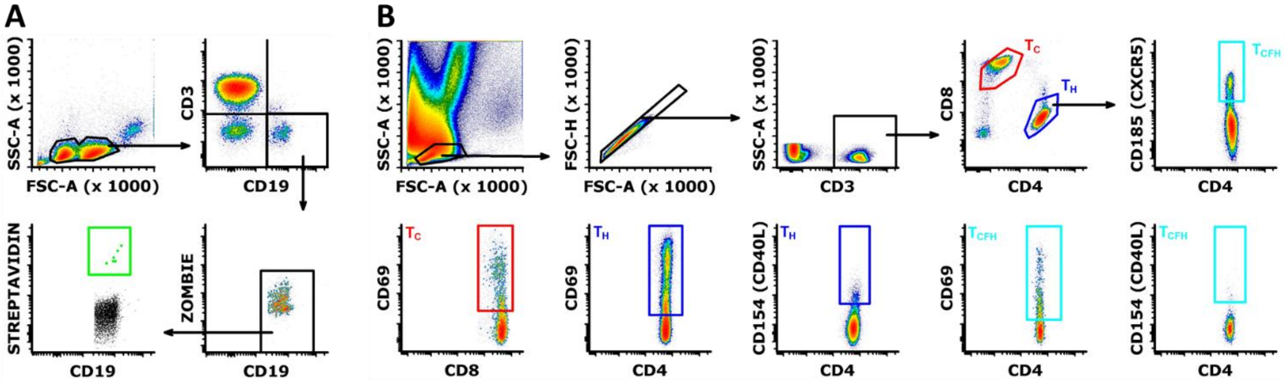


**Supplementary Figure 2. Flowcytometric B and T cell profiling. A)** Gating strategy used to detect circulating RBD specific B cells from thawed PBMCs. **B)** Gating strategy used to define the different T cell subsets ($T_H$, $T_C$ and $T_{CFH}$) and to assess T cell activity using both CD69 and CD40L membrane markers from whole blood. Abbreviations: RBD = receptor-binding domain, PBMCs = peripheral blood mononuclear cells.

#### 1.2 Supplementary Tables

**Supplementary Table 1. B and T cell staining panel used for flowcytometric phenotyping.**

| **Marker** | **Fluorochrome** | **Clone** | **Quantity/test** | **Company** |
|---|---|---|---|---|
| ***B cell panel **** | | | | |
| CD3 | FITC | UCHT1 | 1:1000 dilution | Biolegend |
| CD19 | Per-CP/Cy5.5 | HIB19 | 1:400 dilution | Biolegend |
| Streptavidin | PE | NA | 1:1000 dilution | Biolegend |
| Zombie | Aqua | NA | 1:1000 dilution | Biolegend |
| ***T cell panel ***** | | | | |
| CD3 | FITC | UCHT1 | 5 µL / 100 µL WB | Biolegend |
| CD4 | Pacific Blue | SK3 | 5 µL / test on WB | Biolegend |
| CD8 | Brilliant Violet 510 | RPA-T8 | 5 µL / 100 µL WB | Biolegend |
| CXCR5 (CD185) | PE/Cy7 | J252D4 | 5 µL / 100 µL WB | Biolegend |
| CD40L (CD154) | PE | 24-31 | 5 µL / 100 µL WB | Biolegend |
| CD69 | APC | FN50 | 5 µL / 100 µL WB | Biolegend |

* B cell staining panel: quantity/test based on one million PBMCs per tube. ** T cell staining panel: quantity/test values represent those used to perform a whole blood staining. Abbreviations: NA = not applicable, WB = whole blood, PBMC = peripheral blood mononuclear cells.
